# Using Machine Learning to Predict Mortality for COVID-19 Patients on Day Zero in the ICU

**DOI:** 10.1101/2021.02.04.21251131

**Authors:** Elham Jamshidi, Amirhossein Asgary, Nader Tavakoli, Alireza Zali, Hadi Esmaily, Seyed Hamid Jamaldini, Amir Daaee, Amirhesam Babajani, Mohammad Ali Sendani Kashi, Masoud Jamshidi, Sahand Jamal Rahi, Nahal Mansouri

## Abstract

**Rationale:** Given the expanding number of COVID-19 cases and the potential for upcoming waves of infection, there is an urgent need for early prediction of the severity of the disease in intensive care unit (ICU) patients to optimize treatment strategies.

**Objectives:** Early prediction of mortality using machine learning based on typical laboratory results and clinical data registered on the day of ICU admission.

**Methods:** We studied retrospectively 263 COVID-19 ICU patients. To find parameters with the highest predictive values, Kolmogorov-Smirnov and Pearson chi-squared tests were used. Logistic regression and random forest (RF) algorithms were utilized to build classification models. The impact of each marker on the RF model predictions was studied by implementing the local interpretable model-agnostic explanation technique (LIME-SP).

**Results:** Among 66 documented parameters, 15 factors with the highest predictive values were identified as follows: gender, age, blood urea nitrogen (BUN), creatinine, international normalized ratio (INR), albumin, mean corpuscular volume, white blood cell count, segmented neutrophil count, lymphocyte count, red cell distribution width (RDW), and mean cell hemoglobin along with a history of neurological, cardiovascular, and respiratory disorders. Our RF model can predict patients outcomes with a sensitivity of 70% and a specificity of 75%.

**Conclusions:** The most decisive variables in our model were increased levels of BUN, lowered albumin levels, increased creatinine, INR, and RDW along with gender and age. Complete blood count parameters were also crucial for some patients. Considering the importance of early triage decisions, this model can be a useful tool in COVID-19 ICU decision-making.

## Introduction

COVID-19 has currently affected more than 82 million people worldwide and caused more than 1.8 million deaths ^1^. Complications are more common among elderly patients and people with preexisting conditions, and the rate of intensive care unit (ICU) admission is substantially higher in these groups ^2,3^.

ICU admissions rely on the critical care capacity of the health care system. Iran, which is the primary testbed for this study, was one of the first countries hit by COVID-19. The ICU admission rate is around 32% of all hospitalizations, and the ICU mortality rate is about 39% ^4^. With the potential upcoming waves of COVID□19 infections, these numbers are expected to rise, leading to shortages of ICU beds and critical management equipment. There is also the risk of a global shortage of effective medical supplies, making the judicious use of these medications a top priority for healthcare systems.

An individual-based prediction model is essential for tailoring treatment strategies and would aid in expanding our insights into the pathogenesis of COVID-19. A number of risk assessment scores are available to predict the severity of different diseases in ICU patients ^5^. Predictors of the need for intensive respiratory or vasopressor support in patients with COVID-19 and of mortality in COVID-19 patients with pneumonia have been put forth ^6,7^. To date, no general mortality prediction scores have been available for COVID-19 patients presenting to the ICU, irrespective of the patients’ clinical presentation. Additionally, both the existing risk scales rely on parameters measured by health care providers such as blood pressure, respiratory rate, and oxygen saturation, which are subject to human error and operator bias especially under the challenging and stressful conditions when the numbers of COVID-19 patients surge ^8^. Thus, it remains vital to develop more unbiased risk assessment tools that can predict the most likely outcomes for individual patients with COVID-19.

Recent advances in artificial intelligence (AI) technology for disease screening show promise as computer-aided diagnosis and prediction tools ^9–12^.In the era of COVID-19, AI has played an important role in the early diagnosis of infection, contact tracing, and drug and vaccine development ^13^. Thus, AI represents a useful technology for the management of COVID-19 patients with the potential to reduce the mortality rate of this disease. Nevertheless, an AI tool for making standardized and accurate predictions of outcome in COVID-19 patients with severe disease is missing.

Beyond the general benefits of data-driven decision-making, the pandemic has also exposed the need for computational assistance to health care providers, who under the rush of severely ill patients may make mistakes in judgement ^8,14,15^. Stressful conditions and burnout in health care providers can reduce their clinical performance, and a lack of accurate judgment can lead to increased mortality rates ^16,17^. Artificial intelligence can help healthcare professionals determine who needs a critical level of care more precisely. Indeed, the effective use of AI could mitigate the severity of this outbreak.

Here, we propose a personalized machine-learning (ML) method for predicting mortality in COVID-19 patients based on routinely available laboratory and clinical data at the date of ICU admission.

## Methods

### Data Resources

This retrospective study includes data from 263 adult patients with COVID-19 infection confirmed through reverse transcription-polymerase chain reaction (RT-PCR). They were admitted to ICUs at different hospitals in Tehran, Iran between February 19 and May 1, 2020. The study was performed after approval by Iran University of Medical Sciences Ethics Committee (approval ID: IR.IUMS.REC.1399.595)

### Statistical Analysis & Feature Selection

On the day of the ICU admission, 66 parameters were assessed for each patient including 11 demographic characteristics (e.g., age and gender), past medical history and comorbidities (including nine different preexisting conditions), and 55 laboratory biomarkers. These parameters are listed in Table 1. 69% of measurements were reported on the day of admission, 27% were reported one day after, and 4% were reported within two days of ICU admission because of sampling limitations and laboratory practice. We excluded patients whose laboratory data were obtained more than two days after the date of admission to the ICU.

**Table 1:**
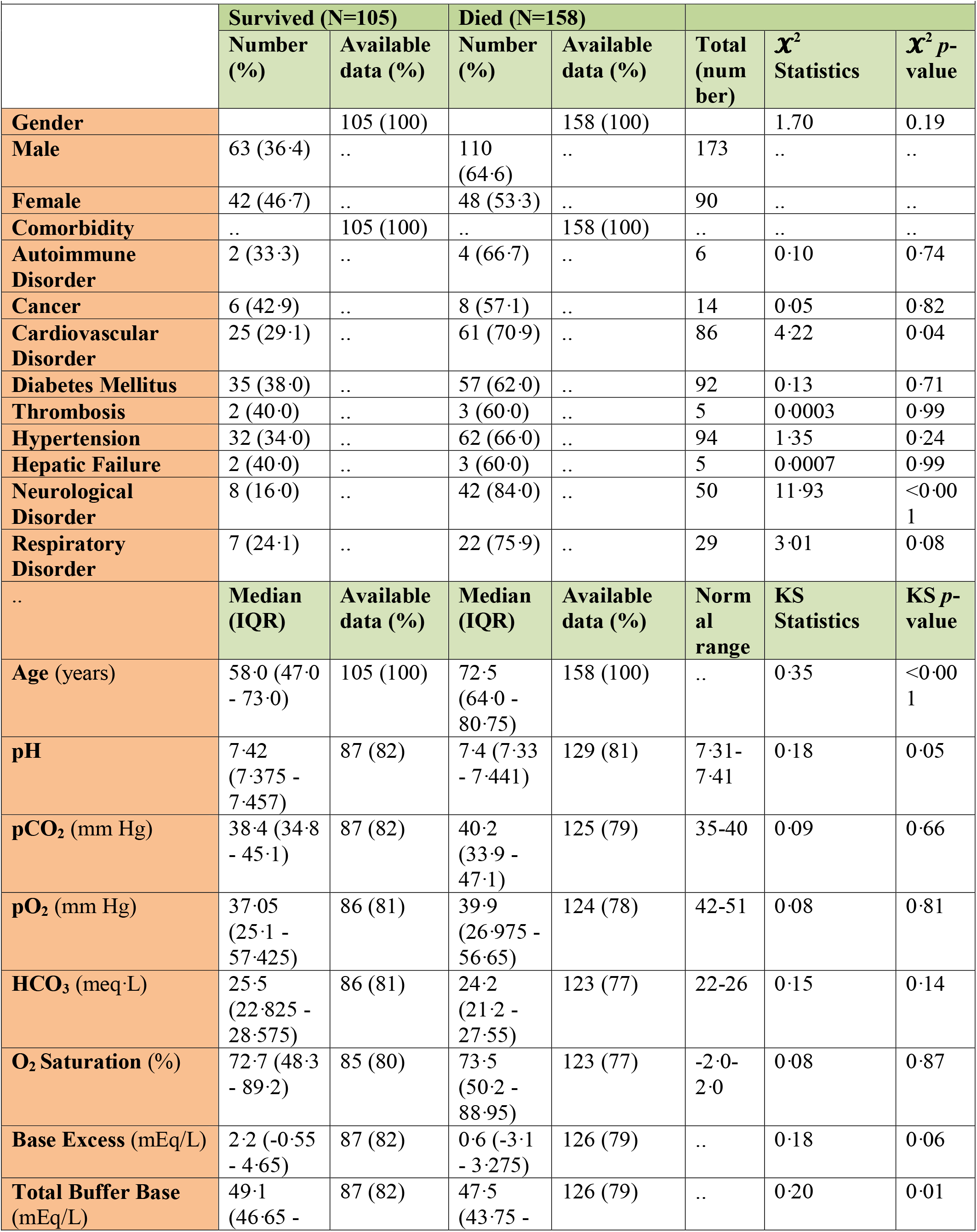
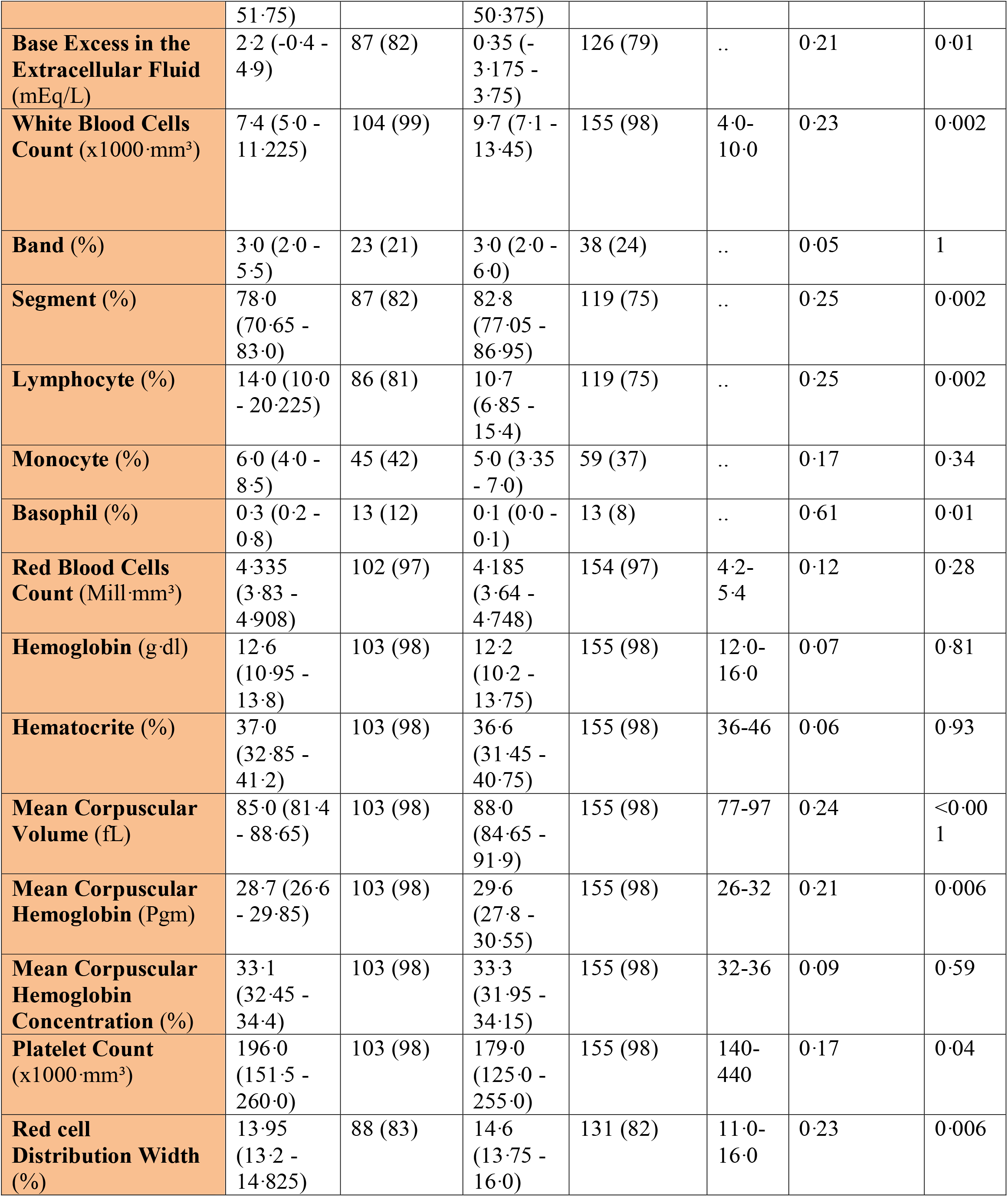
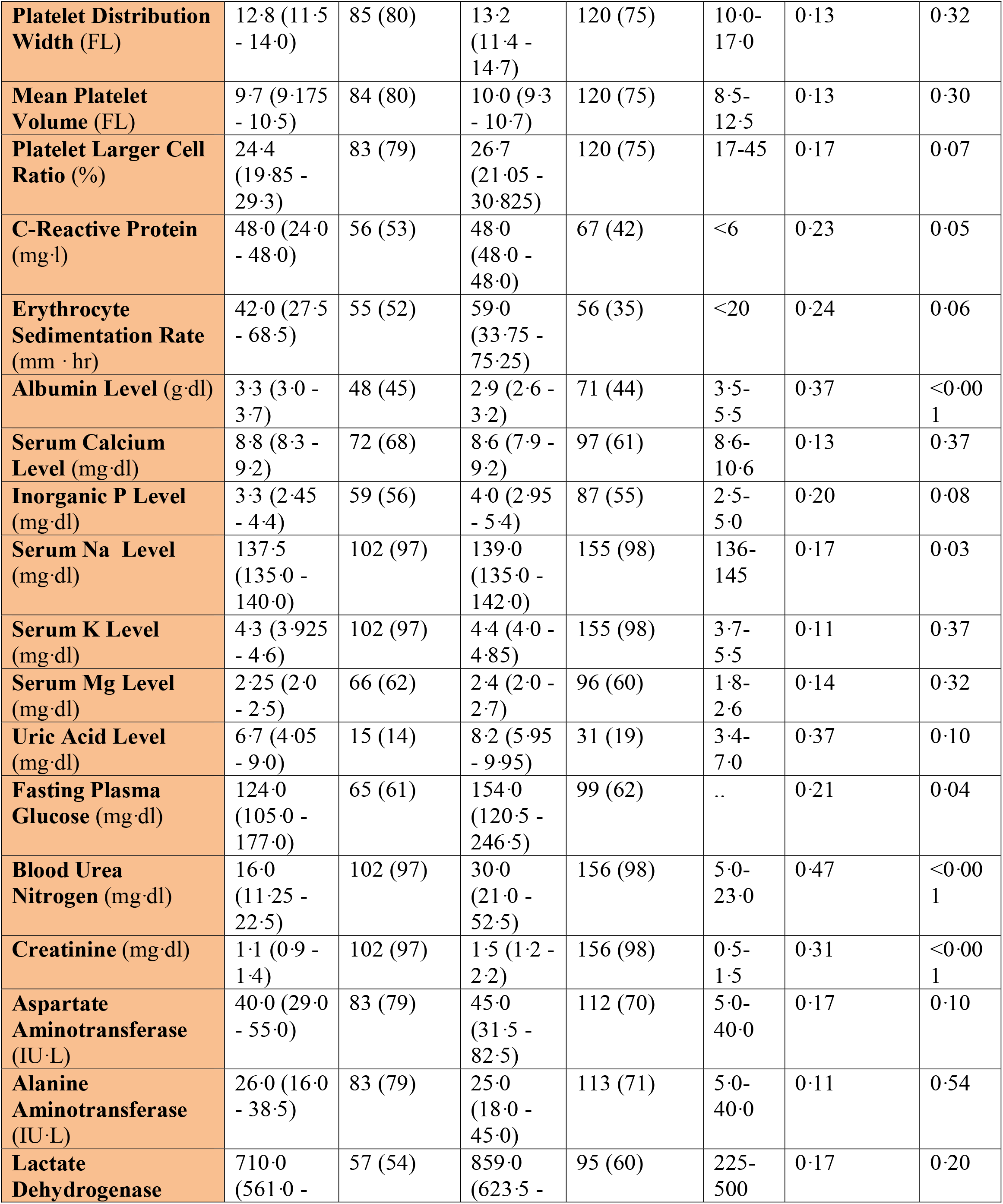
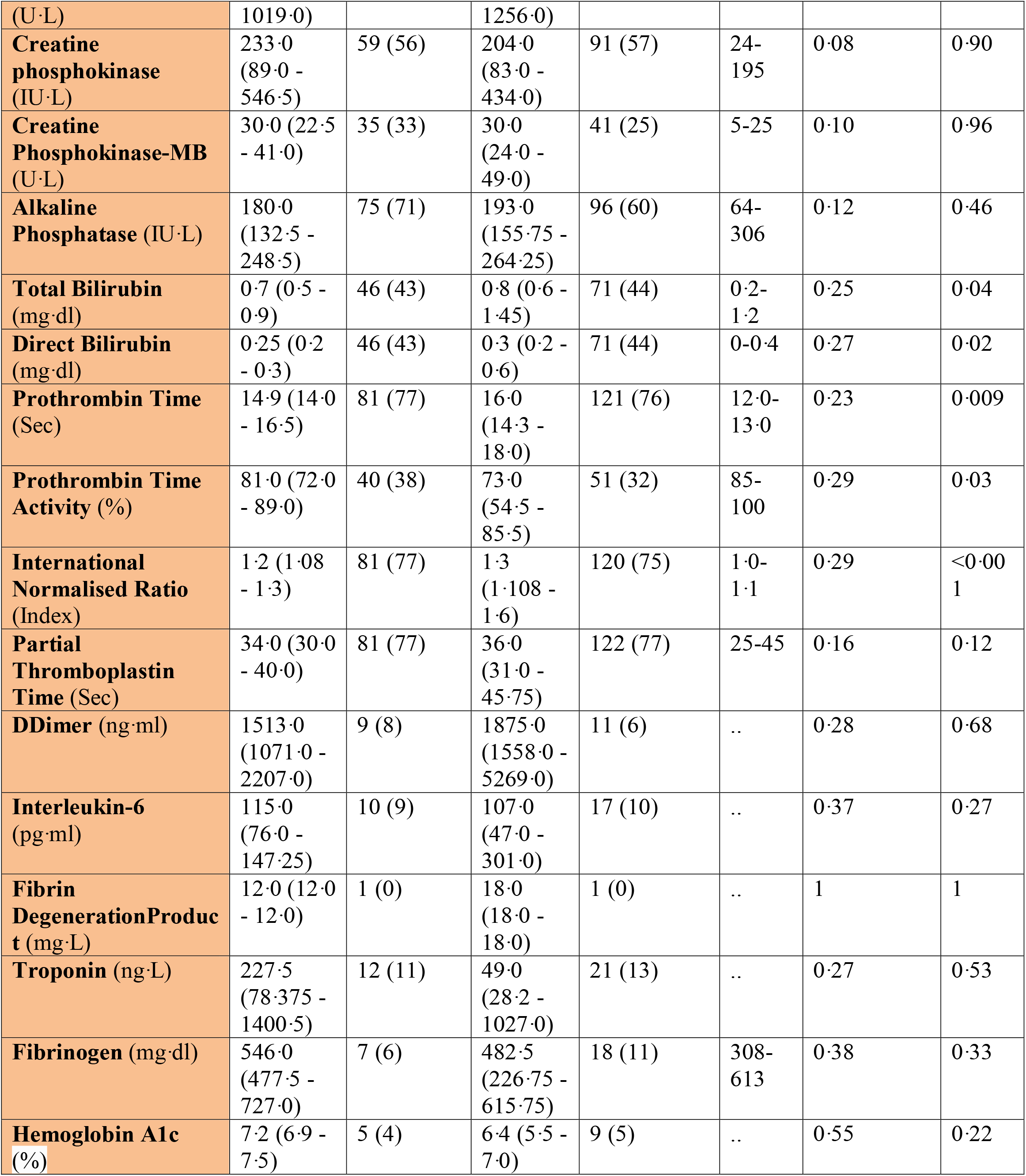
Characteristics of intensive care unit patients with COVID-19 in our data.

The aim was to predict patient survival. For the selection of parameters with the highest predictive value, under the null hypothesis of distributions being the same between the two groups, the two-sample Kolmogorov-Smirnov test (KS), shown in Supplementary Figure 1, was used for numerical parameters (age and laboratory biomarkers), and the Pearson chi-squared test (*X*^2^), shown in Supplementary Figure 2, was used for categorical parameters (e.g., gender and comorbidities). To reduce the model’s feature space dimensionality, only parameters with the highest predictive values were used in the modeling, leading to more robustness and generalizability of the model.^18^

### Data Preprocessing

Data processing was carried out in four steps: First, because of incomplete laboratory data and in order to reduce difficulties associated with missing values, 235 patients having data for at least seven out of ten biomarkers were selected. Second, samples were randomly separated into 10 independent sets with stratification over outcomes for 10-fold cross-validation to ensure the generalizability of the models ^19^. Of the 10 subsets, a single subset was retained as a validation set for model testing and the remaining nine subsets were used as training data. The cross-validation process was then iterated ten times with each of the 10 subsets being used as the validation data exactly once. Third, numerical parameters were standardized by scaling the features to mean zero and unit variance. Lastly, missing biomarker values were imputed using the k-nearest neighbor (k-NN) algorithm, and a binary indicator of missingness for each biomarker was added to the dataset ^18,20^. Standardization and imputation were performed separately on each cross-validation iteration by using training set samples.

### Machine Learning Model

Logistic regression (LR) and random forest (RF) methods were used to build classification models using the Python scikit-learn package ^21^. The performance of each method on training and validation sets in each cross-validation iteration was compared using a receiver operating characteristic curve (ROC), which is shown in Figure 1. To prevent overfitting in the training process, the LR model was trained with an L2 regularization factor equal to one, and the RF was forced to hold more than 10% of samples in each of its terminal leaves ^22,23^. To find the most influential parameters in the LR model prediction, we used regression coefficients, which is shown in the Supplementary Material. Using the local interpretable model-agnostic explanation submodular-pick (LIME-SP) method, we identified different patterns among the whole feature space in the RF model ^24^. The LIME-SP method can interpret the model’s predictions in different parts of the feature space by modeling a subset of model predictions in the feature space around the sample with the help of linear models that are more interpretable. In our study, LIME-SP was performed on 100 random samples to find six submodules with the most disparity in their selected markers, as shown in Figure 2. To identify meaningful clinical differences between patients, seven parameters with the highest predictive values were derived from each submodule.

**Fig. 1.**
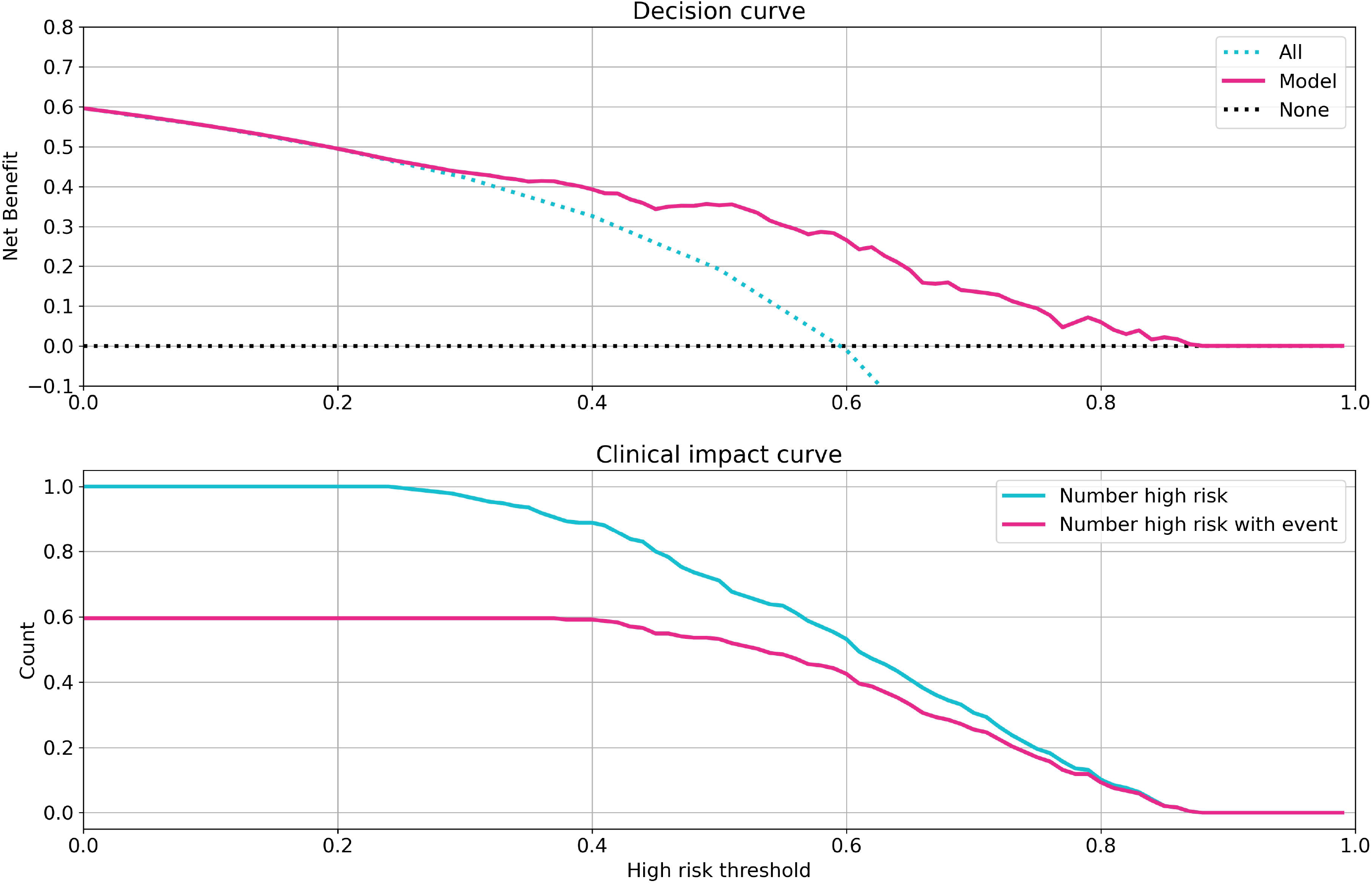
Investigation of model performance. Receiver operating characteristic curve of logistic regression (top) and random forest (bottom) models for training and validation sets for each cross-validation iteration and corresponding area under curves (AUC)s. The random forest model shows superior performance on both training and validation sets. The random forest model predicts patient outcomes with a 70% sensitivity and 75% specificity.

**Fig. 2.**
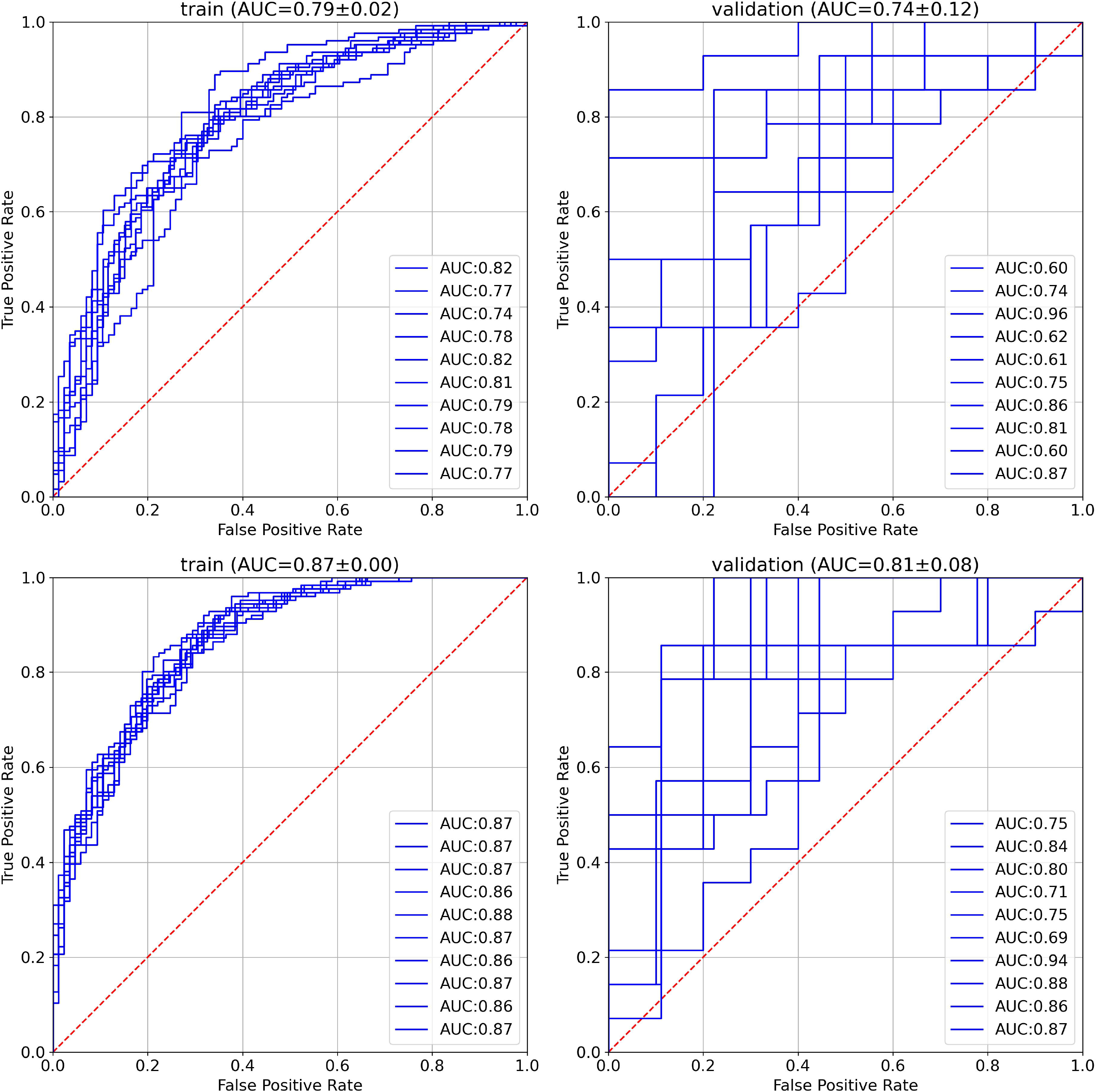
Feature importance in random forest model. The importance of the random forest features using local interpretable model-agnostic explanation submodular-pick with six submodules. Negative values (blue) indicate favorable parameters suggesting a better prognosis, and positive values (red) indicate unfavorable parameters suggesting a worse prognosis.

## Results

The median age of patients was 69 years with an interquartile range (IQR) of 54-78. The minimum and maximum ages were 20 and 98 years, respectively. One hundred fifty-three patients (65.1%) were men, and 82 (34.9%) were women. One hundred five (39.9%) were discharged from the ICU after recovery and 158 (60.1%) patients lost their lives. The most frequent comorbidities among the patients were hypertension, diabetes, and cardiovascular disorders in 94, 92, and 86 patients, respectively. Among the 158 deceased patients, neurological disorders were the most prevalent comorbidity (42 patients, 84%). The statistical analysis and the availability of each parameter in our dataset are summarized in Table 1.

In the RF model, the optimum point between overfitting and efficiency was found by selecting 10 laboratory biomarkers out of 55 with the lowest KS *p*-values and three out of nine comorbidities with the lowest *X*^2^ *p*-values, besides demographic characteristics.

The selected numerical parameters for modeling were as follows: age, blood urea nitrogen (BUN), serum creatinine level (Cr), international normalized ratio (INR), serum albumin, mean corpuscular volume (MCV), red cell distribution width (RDW), mean corpuscular hemoglobin (MCH), white blood cell count (WBC), segmented neutrophil count, and lymphocyte count. In addition, selected categorical parameters were gender and a history of neurological, respiratory, and cardiovascular diseases. The distributions of selected numerical (age and biomarkers) and categorical (gender and preexisting conditions) variables are shown in Supplementary Figure 4 and Figure 5, respectively.

Based on the ROC curves of the models (Figure 1), the RF model outperformed the LR model in both training and validation sets and had superior efficiency. The better performance of RF could be explained by the complexity of the effects of COVID-19 and the varied etiologies for the deterioration of COVID-19 patients due to which the nonlinear characteristics of the RF model made it a more suitable option for predictions than the linear LR model. The RF model can predict a patient’s outcome with a sensitivity of 70% and a specificity of 75%, while the sensitivity for the LR model was 65% and the specificity was 70%.

By using the LIME technique, variables that provide the most information on the probability of each patient’s death were identified.

Among the six submodules identified with the highest disparity among 100 patients, albumin, BUN, and RDW were present in five of them. Age, MCH, and creatinine were present in four of the abovementioned submodules. This points out the importance of these measurements in the recorded parameters. Additionally, BUN (in three of these submodules), RDW (in two submodules), and age (in one submodule), were the most decisive ones.

This model could predict a patient’s outcome reliably (AUC between 80 to 85) over a 15-day period, as shown in Figure 3. The mortality rate was highest between zero and four days. Given that the model was designed for first-day ICU admissions, moving away from this day reduced the accuracy of the predictions and the efficacy of the LIME method for clinical interventions, as expected.

**Fig. 3.**
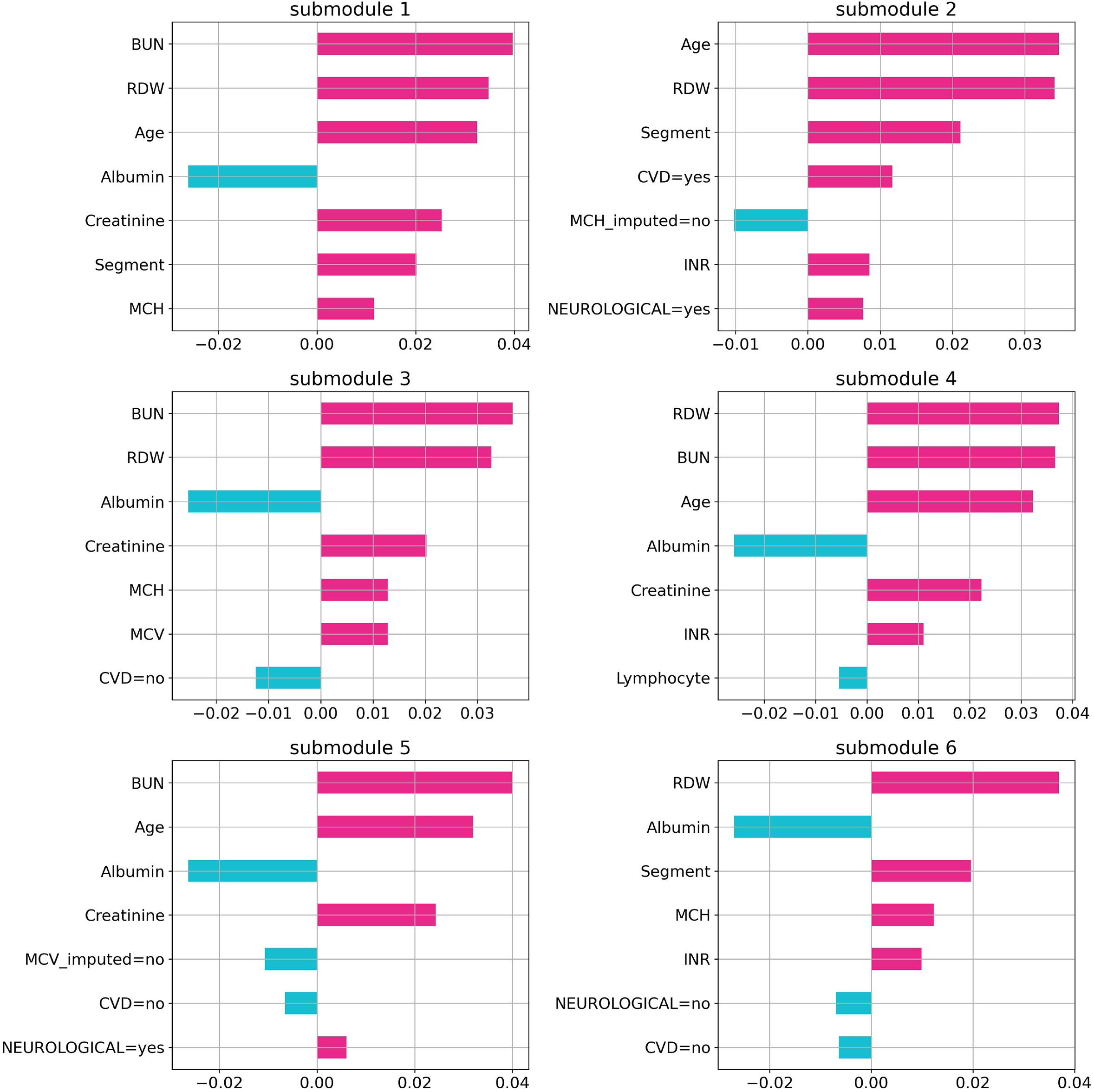
The relation between prediction horizon and performance. Distribution of days between intensive care unit admission and outcome (bars) and corresponding random forest model’s area under the receiver operating characteristic curve scores for each bin (red line). Our model has the best performance to predict outcomes in a 15-day period.

To evaluate the clinical capability of the model, the decision curve (DC), and the clinical impact curve (CIC) were investigated ^25^. The DC framework measures the clinical “net benefit” for the prediction model relative to the current treatment strategy for all or no patients. Net benefit is measured over a spectrum of threshold probabilities, defined as the minimum disease risk at which further intervention is required. Based on the DC, CIC, and on the assumption of the same interventions for high-risk patients, our model indicated a superior or equal net benefit within a wide range of risk thresholds and patient outcomes, as shown in Figure 4.

**Fig. 4.**
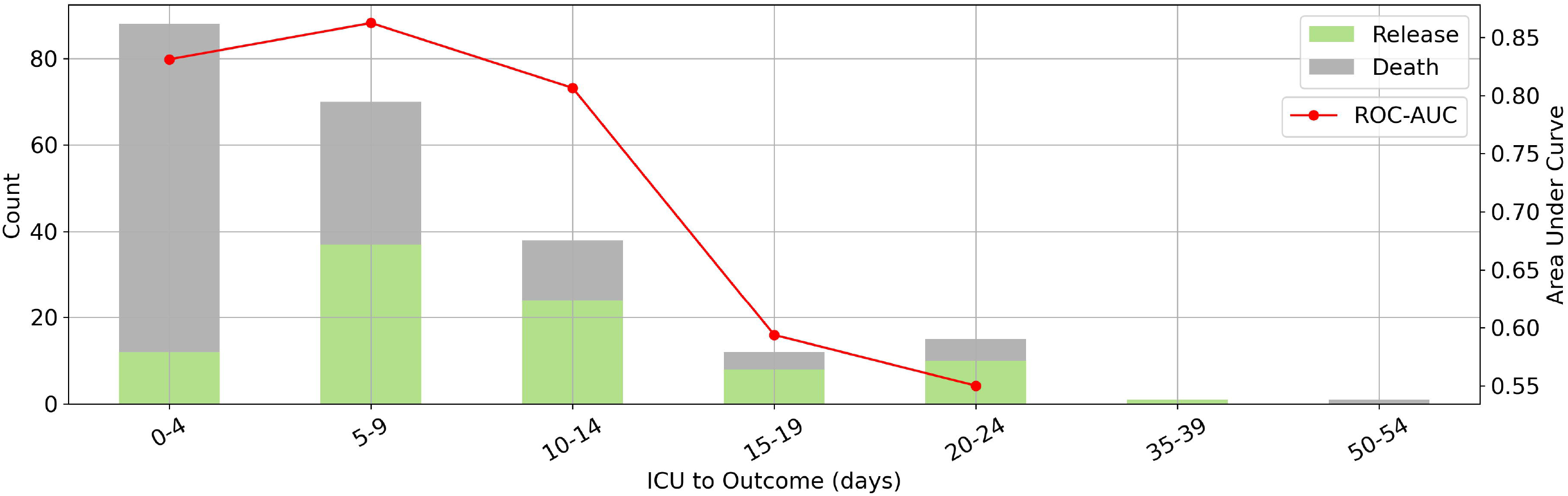
Investigation of clinical impacts and benefits of the model. Decision curve (top) and clinical impact curve (bottom) of the random forest model. The decision curve compares the net benefits of an intervention in three scenarios: for all patients, for no patients, and for high-risk patients based on the model prediction. The clinical impact curve compares the number of patients classified as high risk and the number of patients with a poor prognosis who were classified as high risk, for each high-risk threshold.

## Discussion

The aim of this study was developing an interpretable ML model to predict the mortality rate of COVID-19 patients at the time of admission to the ICU. To the best of our knowledge, this is the first study to develop a predictive model of mortality in patients with severe COVID-19 infection at such an early stage using routine laboratory results and demographic characteristics.

The most decisive parameters based on the two-sample KS test were, in decreasing order of importance, increased BUN, Cr, INR, MCV, WBC, segmented neutrophils count, RDW, MCH, and decreased albumin and lymphocyte levels. Moreover, based on a *X*^2^ test, age, gender, and a history of neurological, cardiovascular, and respiratory disorders were identified as parameters with high predictive values.

A number of studies have investigated the risk factors affecting COVID-19 infections. Elevated inflammatory cytokines such as interleukin-6 (IL-6), granulocyte colony-stimulating factor (G-CSF), interferon gamma-induced protein 10 (IP-10), and interferon (IFN)-γhave been suggested as poor prognostic factors for COVID-19 patients ^26–28^. These markers, however, are not usually used as predictors of the severity of disease in clinical practice. Although using these cytokines in modeling may enable a more accurate prediction of the severity of COVID-19 infection, doing so impedes the model’s clinical application, as most of the cytokines are not routinely checked at presentation to the ICU. In contrast, all 10 laboratory biomarkers identified in our model are commonly measured and are available to most clinical laboratories. Thus, the DC and CIC analyses indicated the notable clinical benefit of our model especially in a situation characterized by resource scarcity.

Only patients who had at least seven of the 10 selected biomarkers have been included in the training phase of the modeling and missing parameters were imputed using k-NN based on the data. As can be seen in the models’ ROC curve, the RF algorithm outperformed LR in predicting the outcome. This difference is mainly due to the non-linear correlation between variables, manifesting the complexity of the problem.

The application of the LIME-SM method allowed us to determine a patient-specific marker set that each patient’s prognosis is based on. This technique explains the predictions by perturbing the input of data samples and evaluating the effects. The output of LIME is a list of features, reflecting each feature’s contribution to a given prediction. Understanding the ‘reasoning’ of the ML model is crucial for increasing physicians’ confidence in selecting treatments based on the prognosis scores. Using the LIME method, the significance of variables with high predictive value was determined for each prediction made for an individual. The evaluation of the variables in the individual’s personalized prediction can lead to supportive measures and help determine treatment strategies according to the interpretation of the individual prognosis.

As severe COVID-19 may result from various underlying etiologies, our model can help categorize patients into groups with distinct clinical prognosis, thus allowing personalized treatments. In addition to targeted therapies, the differentiation between patients may reveal disease mechanisms that coincide or that occur under specific preexisting conditions. Future cohort studies could explore these assumptions with increased sample sizes.

In this study, hypoalbuminemia and renal function were identified as the main factors with high predictive values for the model. These findings are in agreement with recent results showing that hypoalbuminemia is an indicator of poor prognosis for COVID-19 patients ^29^. It is well documented that endogenous albumin is the primary extracellular molecule responsible for regulating the plasma redox state among plasma antioxidants ^30^. Moreover, it has been shown that albumin downregulates the expression of the angiotensin-converting enzyme 2 (ACE2) which may explain the association of hypoalbuminemia with severe COVID-19 ^31^. Intravenous albumin therapy has been shown to improve multiple organ functions ^32^. Therefore, early treatment with human albumin in severe cases of COVID-19 patients before the drop in albumin levels might have positive outcomes and needs to be further investigated.

Furthermore, increased levels of BUN and Cr are observed in our study, which is an indication of kidney damage. An abrupt loss of kidney function in COVID-19 is strongly associated with increased mortality and morbidity ^33^. There are multiple mechanisms supporting this association ^34,35^.

One of the findings of this study is the identification of RDW (a measure of the variability of the sizes of RBCs) as an influential parameter. This result is in line with recently published reports^36^. Elevated RDW, known as anisocytosis, reflects a higher heterogeneity in erythrocyte sizes caused by erythrocyte maturation and degradation abnormalities. Several studies have found that elevated RDW is associated with inflammatory markers in the blood such as IL-6, tumor necrosis factor-α, and CRP, which is common in severely ill Covid-19 patients ^37^. These inflammatory markers could disrupt the erythropoiesis by directly suppressing erythroid precursors, promoting apoptosis of precursor cells, and reducing the bioavailability of iron for hemoglobin synthesis.

Yan et al. recently identified LDH, lymphocyte, and high-sensitivity C-reactive protein (hs-CRP) as predictors of mortality in COVID-19 patients during their hospitalization. The blood results of hospitalized patients on different days after the initial ICU admission were used for their model^38^. Since our goal was the prediction of mortality risk as early as possible for ICU patients, this limited us to using only the laboratory results on day zero, in contrast. For patients with severe COVID-19 infection, early decision making is critical for successful clinical management. Additionally, laboratory results from other days may not always become available. We also identified lymphocyte count as a predictor of mortality, as in the previous study; however, CRP levels and LDH did not reach statistical significance.

Although IL-6 has been found to be a good predictor of disease severity by other studies, it did not reach statistical significance in our model ^39^. IL-6 had a considerable KS statistical value, but because of the high number of missing values, its *p*-value was not significant compared to other markers. The fact that IL-6 is not always measured upon ICU admission is precisely why it is not suitable for our purposes.

The missingness indicator of some markers in both LR and RF models has an impact on the predictions based on the regression coefficient and LIME, which can be the result of the model compensating for the imputation error. However, the missingness indicator may also indicate the existence of bias in biomarker reporting ^40^. Such biases (e.g., sampling bias) are an inevitable part of retrospective studies. They can be addressed using domain-adaptation techniques such as correlation alignment (CORAL) in future studies using additional data ^41,42^. Another limitation of this study may be the lack of an objective criterion for ICU admission. Moreover, different treatment strategies can change the survival outcome for patients who may have had similar profiles when admitted to the ICU. In future studies, the accuracy of this model may be further improved by adding chest imaging data and by using a larger dataset. Possible targets for our ML framework include the prediction of other crucial information such as the patients’ need for mechanical ventilation, the occurrence of cytokine release syndrome, the severity of acute respiratory disease syndrome, the cause of death, and the right treatment strategy.

In conclusion, we evaluated 66 parameters in COVID-19 patients at the time of ICU admission. Of those parameters, 15 metrics with the highest prediction values were identified: gender, age, BUN, Cr, INR, albumin, MCV, RDW, MCH, WBC, segmented neutrophil count, lymphocyte count, and past medical history of neurological, respiratory, and cardiovascular disorders. In addition, by using the LIME-SP method, we identified different submodules clarifying distinct clinical manifestations of severe COVID-19. The ML model trained in this study could help clinicians determine rapidly which patients are likely to have worse outcomes, and given the limited resources and reliance on supportive care allow physicians to make more informed decisions.

## Data Availability

The data that support the findings of this study are available from the corresponding authors upon request.

## Abbreviations list

ACE2: Angiotensin-Converting Enzyme 2
AI: Artificial Intelligence
BUN: Blood Urea Nitrogen
COVID-19: coronavirus disease of 2019
CIC: clinical impact curve
Cr: Creatinine
CRP: C reactive protein
DC: decision curve
ICU: Intensive care unit
INR: International Normalized Ratio
IFN: interferon
IL-6: Interleukin 6
IQR: interquartile range
KS: Kolmogorov-Smirnov
LR: Logistics regression
LIME: local interpretable model-agnostic explanation
LIME-SP: local interpretable model-agnostic explanation submodular-pick
ML: Machine learning
MCH: mean corpuscular hemoglobin
MCV: mean corpuscular volume
RF: Random forest
RDW: Red blood cell distribution width
ROC: receiver operating characteristic curve
RT-PCR: reverse transcription-polymerase chain reaction
WBC: white blood cells count

## List of figures and legends

**Supplementary Fig. 1. Result of the two-sample Kolmogorov-Smirnov test for all numerical parameters**. 11 out of 56 numerical features with least Kolmogorov-Smirnov *p*-values are used for modeling.

**Supplementary Fig. 2. Result of** *X***2 test for all categorical parameters**. Four out of 56 categorical features with least Kolmogorov-Smirnov *p*-values were used for modeling.

**Supplementary Fig. 3. Regression coefficients of the logistic model**. Features with higher coefficient absolute value were more crucial in the logistic model prediction.

**Supplementary Fig. 4. Distribution of selected numerical variables**. Comparison of categorical variables between released (green) and dead (red) patients.

**Supplementary Fig. 5. Distribution of selected categorical variables**. Comparison of categorical variables between released and dead patients

