## Supplementary Material for "Using Machine Learning to Predict Mortality for COVID-19 Patients on Day Zero in the ICU"

**Supplementary Fig. 1. Result of two-sample Kolmogorov-Smirnov test for all numerical parameters.**

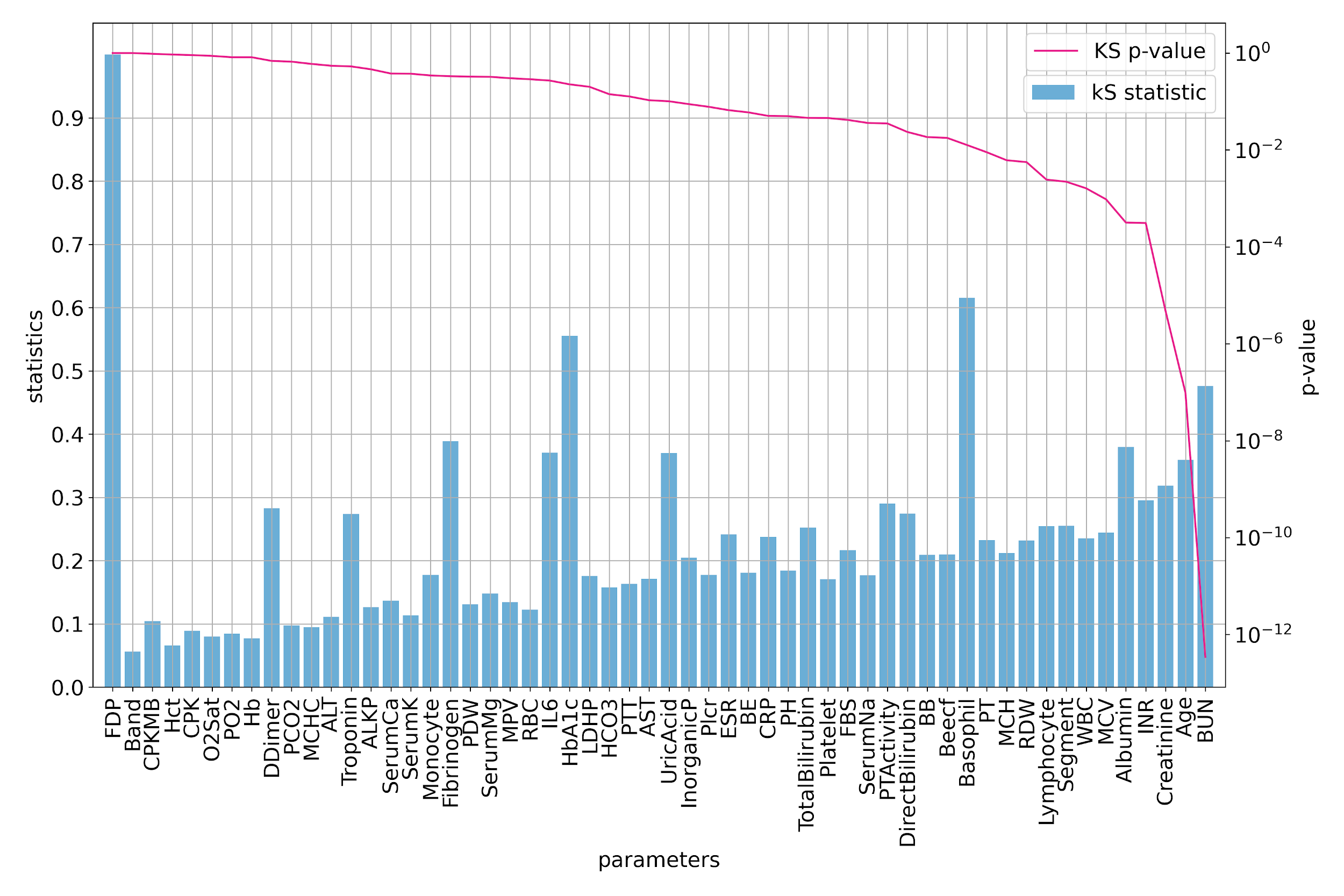

Supplementary Fig. 1. 11 out of 56 numerical features with least Kolmogorov-Smirnov P-values are used for modeling.

**Supplementary Fig. 2.** **Result of 𝓧^2^ test for all categorical parameters**.

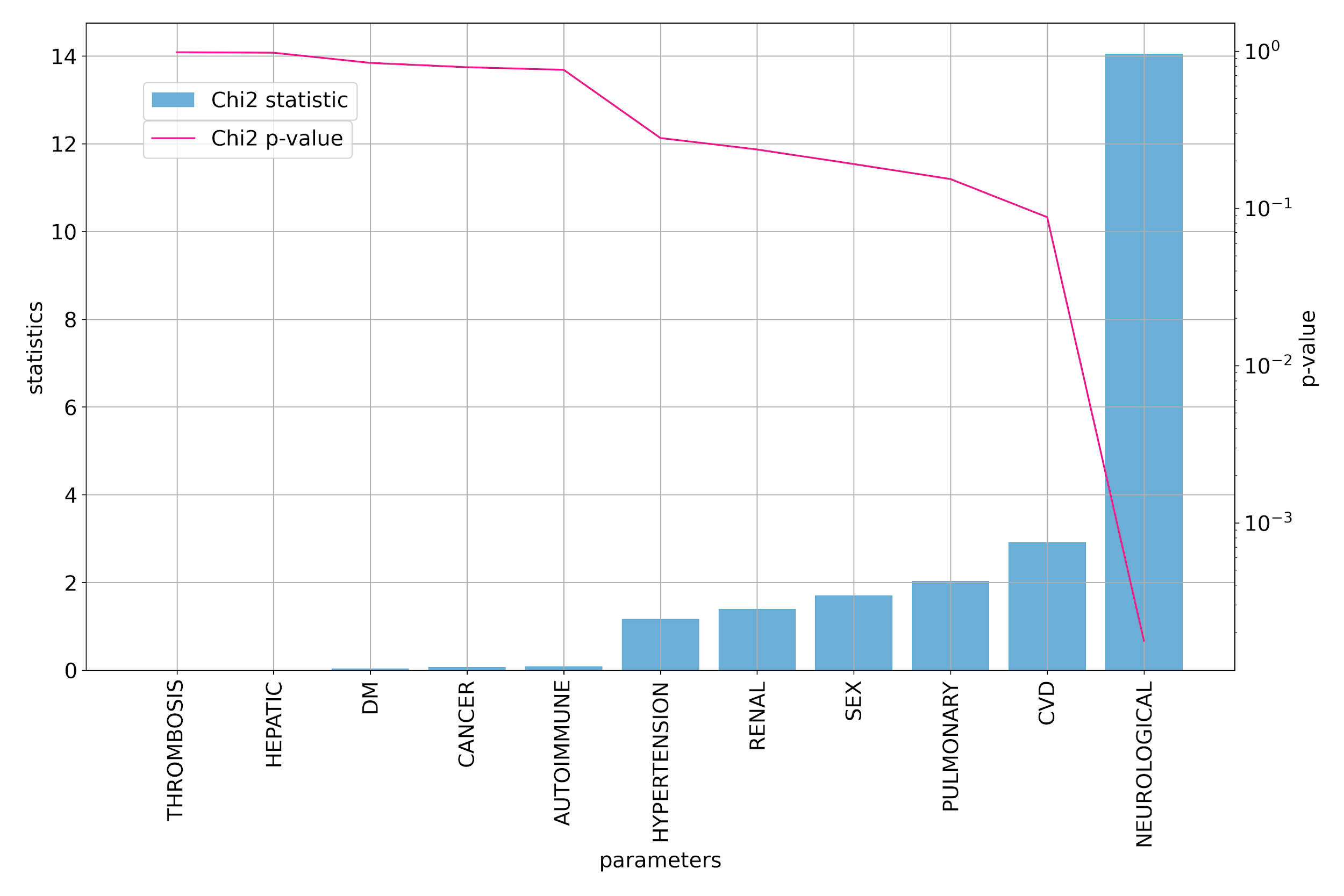

Supplementary Fig. 2. four out of 56 categorical features with least Kolmogorov-Smirnov P-values are used for modeling.

**Supplementary Fig. 3. Regression coefficients of the logistic model.**

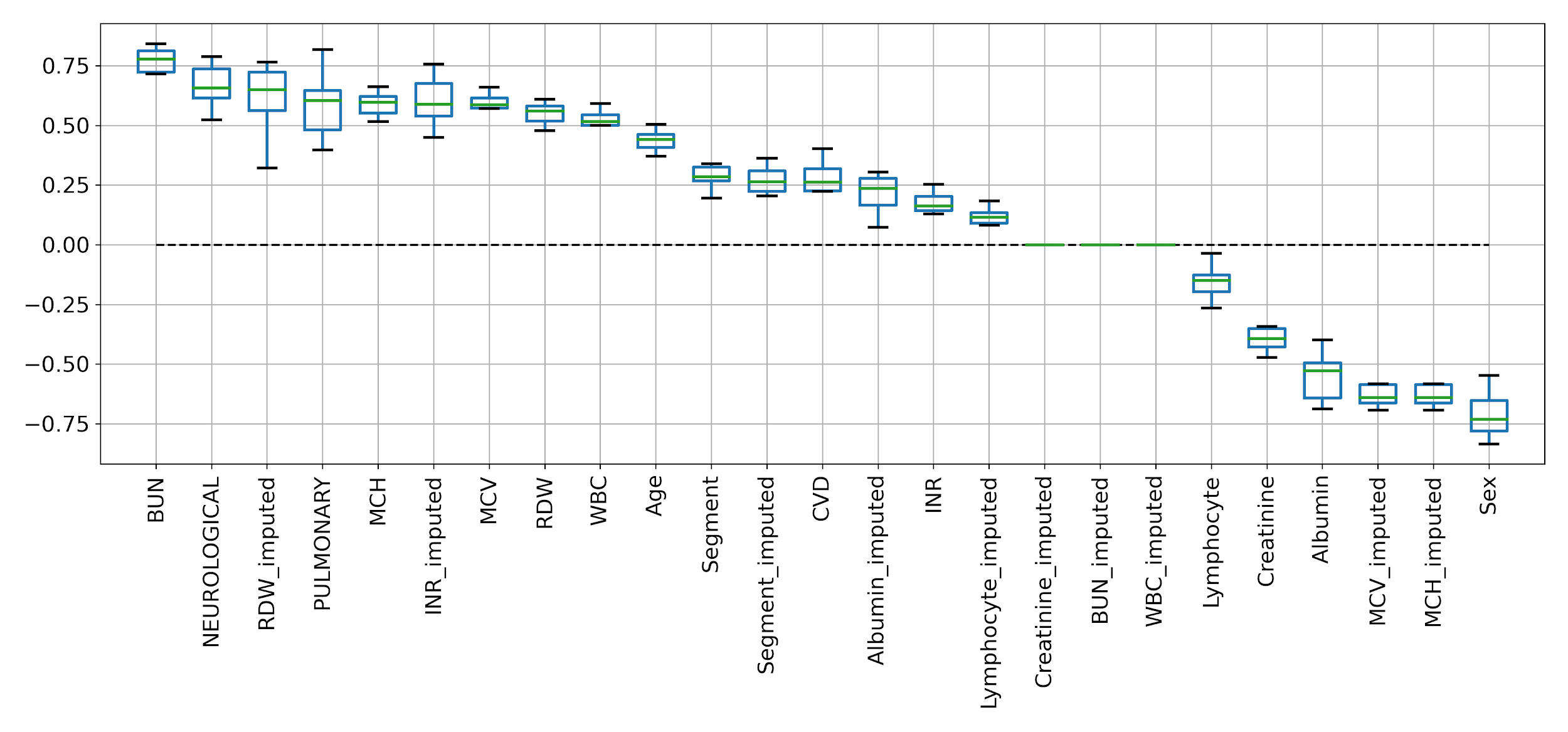

Supplementary Fig. 3. Features with higher coefficient absolute value are more crucial in the logistic model prediction.

**Supplementary Fig. 4. Distribution of selected numerical variables.**

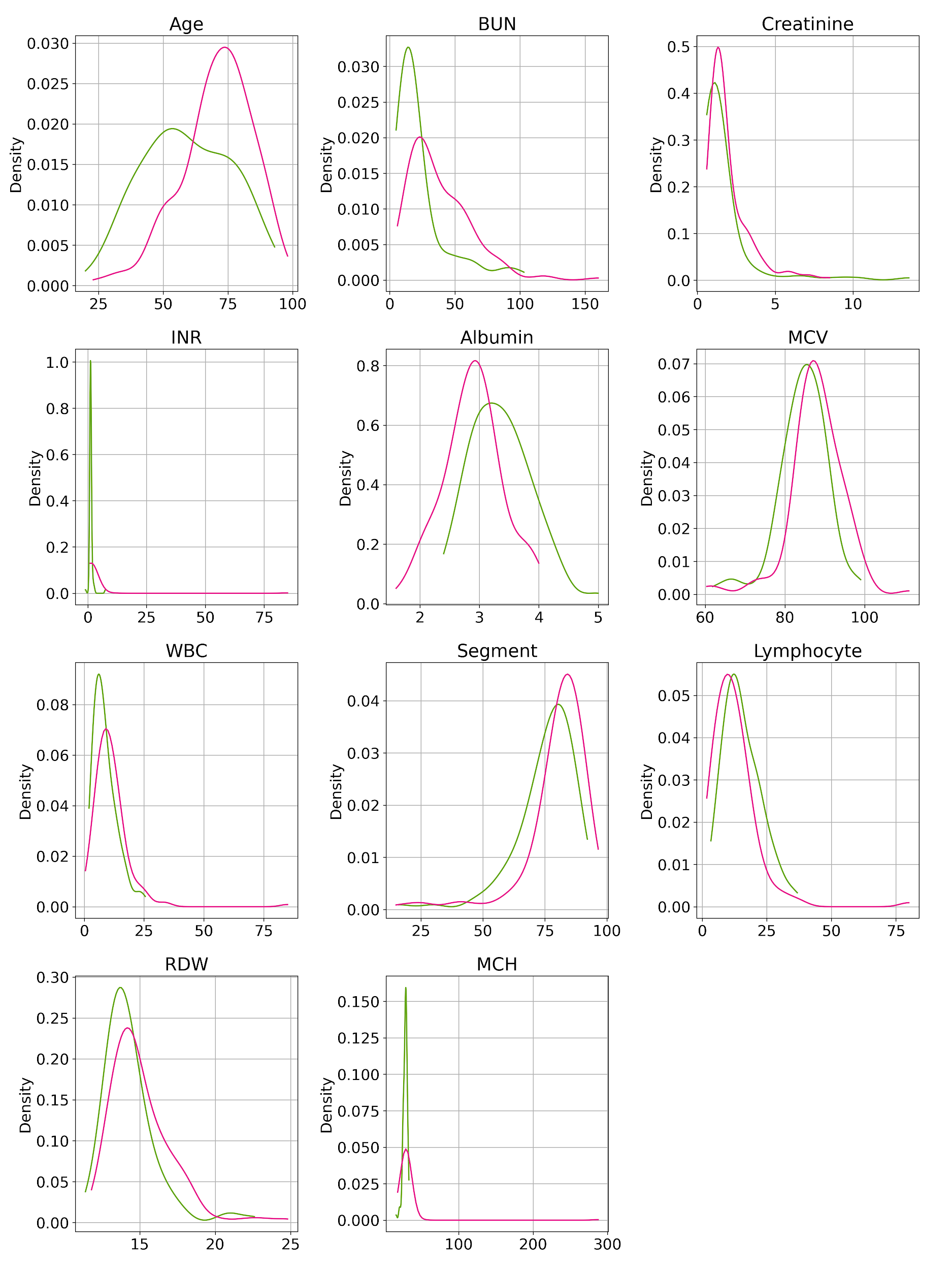

Supplementary Fig. 4. Comparison of categorical variables between released (green) on dead (red) patients.

**Supplementary Fig. 5.** **Distribution of selected categorical variable.**

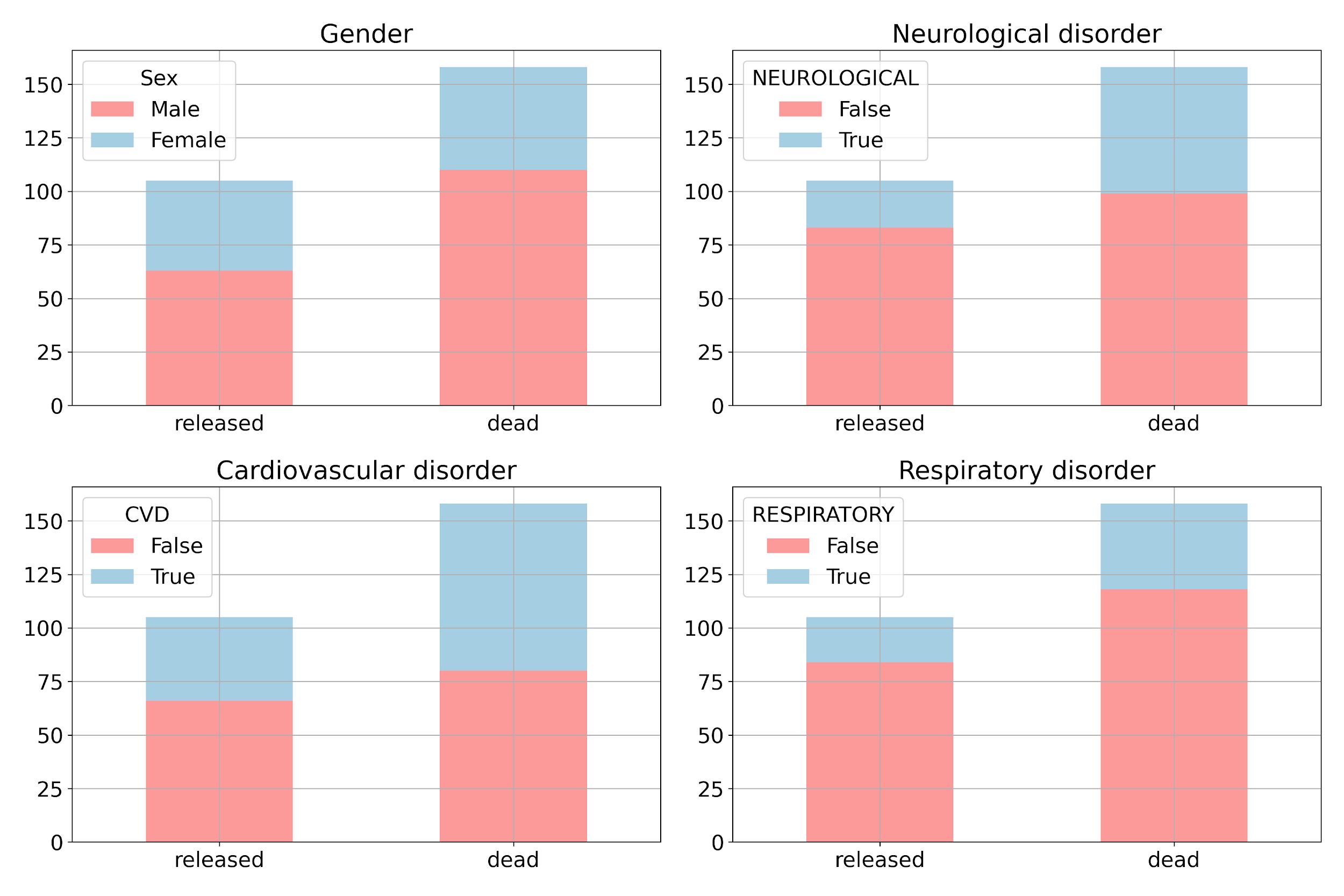

Supplementary Fig. 5. Comparison of categorical variables between released and dead patients.
